# PREVALENCE AND FACTORS ASSOCIATED WITH ASYMPTOMATIC HYPOGLYCEMIA AMONG PRETERM NEWBORNS IN DAR ES SALAAM REGIONAL REFERRAL HOSPITALS, TANZANIA: A CROSS SECTIONAL ANALYTICAL STUDY PROTOCOL

**DOI:** 10.1101/2022.10.28.22281650

**Authors:** Shani Shamsi Salum, Florence Salvatory Kalabamu, Maulidi Rashid Fataki, Salha Ally Omary, Ummulkheir Hamid Mohamed, Hillary Abdillah Kizwi, Kelvin Melkizedeck Leshabari

## Abstract

**Background:** Hypoglycemia is the most common metabolic abnormality in newborns. It is still unclear whether the condition is truly pathological, especially if it occurs transiently during the 1st 24 hours of birth in an asymptomatic phase. Besides, there is hardly any data on the burden of asymptomatic neonatal hypoglycemia and its associated factors among preterm newborns in Africa.

**Aim:** To assess the prevalence and associated factors of asymptomatic neonatal hypoglycemia among preterm newborns in a typical African settings.

**Design and Methods:** We plan to conduct a quick, cross-sectional analytical hospital-based survey at all public regional referral hospitals in Dar es Salaam, Tanzania. We will consider all preterm newborns delivered in the specified settings between June 2022 and December 2022. Our study population will be all preterm newborns delivered at Dar es Salaam public regional referral hospitals. Our target population will be all preterm newborns with asymptomatic hypoglycemia. All newborns with clinically detected congenital anomalies and those who will be delivered at home but brought to the facilities for care will thus be excluded from the study. Our primary outcome measure will be neonatal RBG < 2.6 mmol/L without any symptoms associated with hypoglycemia. Maternal, fetal and early neonatal (> 6 hours but within 24 hours post-delivery) factors will be logistically regressed against the outcome variable after appropriate model validation. Unless otherwise stated, an α-level of 5% will be used as a limit of type I error in findings. Written informed consent will be obtained from mothers of each newborn prior to inclusion into the study.

**Main Outcome measure:** Prevalence of asymptomatic hypoglycemia among preterm newborns in Dar es Salaam hospitals.

**Relevance of the findings to science, policy & practice:** Current clinical practice does not provide evidence for routine glycaemic screening among preterm newborns asymptomatic for hypoglycemia. The study will have a potential to assess stata of preterm newborn with asymptomatic hypoglycemia

## Introduction

Hypoglycemia, or low blood glucose concentration among newborns soon after birth, has been reported as the commonest *metabolic paradox* globally. [1-6] The current clinical management of neonatal hypoglycemia is devoid of a reliable evidence base worldwide. [7, 8] Besides, the current controversy seem to have its root on whether the phenomenon is truly a pathology, especially if it occurs without symptoms among preterm neonates. The most frequent cause of newborn death is preterm birth, which is defined as the birth of a baby prior to actually 37 weeks of pregnancy (9). Around 15 million premature babies are born each year all around the world (10). Preterm birth problems are the major cause of neonatal mortality, accounting for 35% of the world’s 3.1 million deaths each year. (11) They are also the second leading cause of death in children under the age of five, after pneumonia (11). Amidst all the above pahologies, there is a growing body of evidence that links the causes of early neonatal mortality to include neonatal hypoglycemia. (12-14) However, a reliable estimate of prevalence for neonatal hypoglycemia is not known in Tanzania nor do the health system clearly understands factors associated with an even bizzare pattern of *asymptomatic hypoglycemia* among neonates in Tanzania.

On a strict clinical grounds, the finding of *asymptomatic hypoglycemia* to majority of newborn babies has been associated with an altered physiological state of adaptation to extra-uterine life rather than a significant pathology. (15-17) However, when hypoglycemia in newborns is prolonged or recurrent over time, it may result into acute systemic effects, and even long-term neurological sequelae. (18-21) Strangely, a single or two blood glucose levels among newborns have become the grounds for malpractice suits and litigations in US and other Western medical practice; even though the causative relationship between the two phenomena is still considered dubious. (22) Clinically, it is still not possible to define blood glucose thresholds that require medical interventions in every newborn nor is it certain to date on the duration, and level of glycaemia that guarantees damages. (15) Likewise, it is even debatable whether *asymptomatic neonatal hypoglycemia* is a disease state and if so, does warrants any significant medical attention. (23-26) Besides, there are paucity of findings in published literature on the burden and factors associated with *asymptomatic hypoglycemia* in preterm newborn children worldwide. Of the few available evidence in retrievable platforms, almost all have characterised the syndrome among neonates, irrespective of the age of neonates studied. (2-8) It is indeed preterm babies who are *most at risk* of *significant asymptomatic hypoglycemia*. Otherwise, the key evidence on the matter is dominated by studies done among Caucasian newborns. (2-4, 12, 16, 17) The remaining non-Caucasian studies reported elsewhere were from outside Tanzania. (5-7, 13, 14) We thus designed a quick cross-sectional hospital based survey to characterise the burden and factors associated with the syndrome of *asymptomatic hypoglycemia* among preterm neonates at health facilities in Dar es Salaam city, Tanzania.

Globally, there are clear indications that shows how early life stressors are associated with adult health outcomes. (27-32) For instance, Barker and colleagues retrospectively analysed 5654 men born during 1911-1930 from six different districts of Hertfordshire, England for the association between weight at birth and deaths from ischaemic heart disease. (27) They found out deaths from all causes showed a significant downward trend with increasing weight (p<0.001). (27) Furthermore, exclusion of deaths from ischaemic heart disease and chronic obstructive lung disease significantly attenuated the trend. (27) The original report formed the basis of what is currently referred to as *Barker’s hypothesis of fetal programming*. (27) Other studies also reported before, that adult men with ischaemic heart disease to had worse clinical (and socio-economic) states in childhood. (33, 34) On a strict epidemiological sense, it has been proposed that indeed it is timing of childhood adverse conditions that inter alia affects later life course in health. (35) We thus considered preterm neonates as natural surrogates to test whether earlier life environment while in utero have an influence in the development of asymptomatic hypoglycemia. The basis behind our current hypothesis lies on the fact that global prevalence of chronic diseases has raised to pandemic proportions (36-39) Besides, there are clear and reliable evidence for the demographic transition at not only global level; (40) but also in Africa (41) and even more rapidly in Tanzania. (42) We thus believe by identifying the burden and factors associated with insults earlier in life, preventive approaches to late life morbid, and potentially mortal conditions that are at present in pandemic proportion will be eradicated.

Globally, as a result of poor understanding of risk factors and much variability among neonates, even those who are term and healthy, hypoglycemia is not routinely examined in preterm newborns who are not symptomatic. This creates a significant gap in diagnosis, classification as well as prognostic indicators for both morbidity and mortality associated with hypoglycemia in newborn infants. Our study will create an estimate of prevalence and associated factors of hypoglycemia among preterm neonates in Dar es Salaam regional referral facilities. Specifically, we hypothesised that the womb to be more important than the home in development of *asymptomatic hypoglycemia* in preterm newborns as per Sir David Barker’s suggestion back in 1990. (43) From the current proposal, we aim to characterise possible antecedents associated with chronic debilitating conditions in adulthood. Thus, a quick cross-sectional hospital-based analytical study will be conducted at all public regional referral facilities among asymptomatic preterm neonates in Dar es Salaam, Tanzania.

## 2. Materials and Methods

### 2.1 Study aim

To assess the prevalence and factors associated with asymptomatic hypoglycemia among preterm newborns.

### 2.2 Study design

A cross-sectional hospital-based analytical study

### 2.3 Study settings

The research will take place in Dar es Salaam public regional referral hospitals. Specifically, the study will take place at neonatal units of Amana, Mwananyamala and Temeke Regional Referral Hospitals. Dar es Salaam is the business capital of Tanzania. It is a city situated on the East African coast. It is also the 2nd most populated city in the East African region (just behind Kinshasa, DRC). The city’s 5.6 percent average yearly growth rate between the 2002 and 2012 censuses was the highest in the not only in the country and in Africa but among the highest globally. In Dar es Salaam city, there are 3 main referral hospitals feeding directly to the national hospital. Dar es Salaam main hospitals to which the study will take place have average deliveries of 210 per day – ideally seasonal but with no documented data to date.

### 2.4 Study duration

July 2022 to December 2022.

### 2.5 Study population

All preterm newborns in neonatal wards Dar es salaam public regional referral facilities.

### 2.6 Target population

All preterm neonates with asymptomatic hypoglycemia at least 6 hours old and not more than 24 hours after birth.

### 2.7 Sampling method

By default, we plan to recruit all preterm newborn babies delivered at Dar es Salaam public regional referral hospitals after written informed consent of their mothers during the study period.

#### 2.7.1. Inclusion criteria

- Preterm newborn; - Age: 6 hours-24 hours old; - Asymptomatic for hypoglycemia

#### 2.7.2. Exclusion criteria

-Babies with congenital malformation at birth; - Babies delivered at home but referred to the facility

### 2.6 Data collection and analysis

A pre-designed and validated structured questionnaire (main tool) will be used. Reliability and validity testing of the study tool (structured questionnaire) will be assessed using at least 10% of the minimum sample size from at least 2 different private hospital facilities in Dar es Salaam with regional referral status. Specifically, test-retest reliability analysis and construct validity will be the main forms of pretesting the utility of the study tool. Immediately following maternal written informed consent, and given that the participant will be at least 6 hours of age and not more than 24 hour old. Baseline information covering maternal labour activities and events surrounding delivery will be recorded. Specifically, information regarding maternal Hb level, date of birth, level of education, current and previous medical/surgical history, current medication history, known comorbid conditions, obstetric & gynaecologic history, smoking/alcohol/drug abuse stata, current pregnancy type (singleton/multiple), delivery mode (CS/SVD/Assisted delivery) as well as gestational age of a preterm newborn will be assessed using Ballard score chart. For neonatal inputs, growth chart will be used to assess nutritional status (and recorded if it was Small-for-Gestational-Age (SGA), Appropriate-for-Gestational Age (AGA) or Large-for-Gestational-Age-(LGA)), sex of the baby, 1st and 5th minutes Apgar scores, kangaroo care as well as breasfeeding status within the 1st hour after birth. Moreover, neonatal danger signs for sepsis, respiratory distress as well as birth asphyxia will be recorded. Glucometer machine (GoCheck ™) will be used to measure blood glucose of a preterm newborn in mmol/L. Investigators with the assistance of three trained research assistants (intern doctors and nursing officers) will collect data.

Random blood glucose level will be estimated using glucometer machine (GoCheck ™, Microtech Medical Co. Ltd - Hangzhou, China). Using aseptic technique, we will clean the area to be pricked (heel) using alcohol swab and allow to dry before pricking. Blood sample will be obtained from heel of the patient. We will insert glucometer strip into a meter and when the glucometer starts blinking, we will apply the blood sample to the test area on strip. The blood glucose level less than 2.6mmol/l will be considered **hypoglycaemic**. All investigators and the trained research assistants will revise the collected data and triple enter them into SPSS version 23.0 statistical software for further cleaning and analysis.

Principal investigator will check the questionnaire for completeness and accuracy on each day. Completed questionnaires will be numbered and coded before entering in a computer. Cleaning will involve checking for any pertinent errors (e.g. typing errors, coding errors and incomplete data). Exploratory data analysis will be done before main analysis and involve summarising important statistics accordingly as well as identifying any trend and patterns in data structure. Descriptive statistics median (with inter-quartile range) will be used to summarise continuous variables while frequency and percentage will be summary statistics for categorical variables. Main data analysis will employ the usage of generalised linear modelling after appropriate model assumption validation. Specifically, a multivariable logistic regression model will be fitted with the probability of asymptomatic neonatal hypoglycemia as the dependent variable against a series of maternal, fetal and early neonatal variables as independent variables. Wherever possible, a test of interaction terms will also be considered provided that they have a better explanatory power in a linear model. Besides, α-level of 5% will be used as a limit of type 1 error in findings.

Ethical clearance certificate has been sought from the Hubert Kairuki Memorial University’s Institutional Research Ethical Committee as well as the Regional Referral Hospitals Authorities, which includes the Executive Director of Amana Regional Referral Hospital, the Medical Officer in Charge of Mwananyamala Regional Referral Hospital, and the Medical Officer in Charge of Temeke Regional Referral Hospital. Likewise, permission to conduct the study at the specified sites will be sought from municipals’ directors offices for Kinondoni, Ilala and Temeke respectively. Confidentiality of information from each study subjects will be ensured through the use of ID codes to conceal their identity. All responses will be kept private and used for research reasons only and it will be maintained by principle investigator and research assistants during data collection. All recruited participants will have their consent taken by proxy from biological mothers. Specifically, the consent will include the aim of the study, risks and benefits associated with the study participation, a statement on voluntary participation into the study while at the study sites, and that mothers will have all rights to refuse to participate/answer any query in the study tool without jeopardising care received at the institutions as well as a notification for contacting the principal investigator should there be any query/opinion/objection for and against the study participation.

### 3.5 Study limitations

#### Failure to show temporal association between variables

The study has been designed in a cross-sectional fashion, and hence it will not be possible to account for any (if present) temporal association between studied variables.

#### Respondent with incomplete data

There is a possibility that part of participants data will be missing during collection time (e.g. missing information on lab findings) since the study has been designed in a cross-sectional fashion.

#### Record bias

It is also likely that part of the collected information will be erroneous or biased (positively or negatively) altogether by the time of collection from achieved data (e.g. laboratory findings and/or vaccination cards)

## Data Availability

No datasets were generated or analysed during the current study. All relevant data from this study will be made available upon study completion.

## Conflict of interest statement

All authors declare to have no conflict of interest in the preparation and data collection associated with this research topic.

## Investigators’ contributions

**SSS** - Protocol design, funding, pre-testing of the research tool, data collection, future data analysis, initial protocol writing and final approval of the protocol prior to submission process.

**FSK** - Protocol design, future research supervision, future data analysis, final approval of the protocol draft prior to submission process

**MRF** - Protocol design, future research supervision, future data analysis and final approval of the protocol draft prior to submission.

**SAO** - Protocol design, future data analysis and final approval of the protocol draft prior to submission.

**UHM** - Protocol design, future data analysis and final approval of the protocol draft prior to submission.

**HAK** - Protocol design, funding, future data analysis and final approval of the protocol draft prior to submission.

**KL** - Protocol design, pre-testing of the research tool, data analysis, initial protocol draft writing and final approving the protocol draft prior to submission.

## Notes

### Competing Interest Statement

The authors have declared no competing interest.

### Funding Statement

The author(s) received no specific funding for this work

### Author Declarations

Institution's name: Hubert Kairuki Memorial University (HKMU) Clearance certificate number: Ref.No. HKMU/IREC/27.10/151

